# The Golden Hour – Implementing and evaluating a package of interventions to improve newborn care in the first hour of life in a public sector hospital in Zimbabwe

**DOI:** 10.64898/2026.11.06.26367405

**Authors:** Rachel Poffley, Rebekah Soper, David de Lorenzo, Prisca Nyamapfeni, Karlos Madziva, Cyprian Masvikeni, Kevin Meck, Simbarashe Chimhuya, Hannah Gannon, Michelle Heys, Felicity Fitzgerald, Marcia Mangiza, the Neotree Project Team

## Abstract

Reducing neonatal mortality remains a global priority, yet progress has stalled in many low-resource settings. Low-cost interventions such as delayed cord clamping (DCC) and early skin to skin care (ESSC) are known to reduce neonatal morbidity and mortality but implementation remains inconsistent. Prior to project initiation, Sally Mugabe Central Hospital, Zimbabwe’s busiest tertiary maternity unit had no documented DCC or ESSC conducted with practice limited by staffing pressures and infrastructural challenges. We aimed to implement a context-specific “Golden Hour” care bundle to improve newborn care during the first hour after birth.

Using quality improvement (QI) methodology we designed interventions alongside maternity staff, including multidisciplinary training, embedded clinical support, regular feedback, leadership engagement and equipment provision. Real-time progress was tracked using the Neotree digital data tool over 12 months (November 2024-October 2025). The primary outcome was documented DCC and ESSC in eligible infants. An analysis of neonatal mortality among a subset of admitted inborn infants was also carried out.

A total of 207 healthcare workers and 87 student midwives received formal training. Over 12 months, 15,292 eligible infants (81.1%) received DCC and 14,351 (76.1%) received ESSC with sustained implementation between 82-91% (DCC) and 72-85% (ESSC) for the final seven months of the project. Targeted PDSA cycles increased uptake in low birthweight infants and babies born by caesarean section. Logistic regression analysis of infants admitted to the neonatal unit demonstrated a trend to reduction in crude mortality for eligible infants receiving both interventions but this did not reach statistical significance.

A multifaceted, context-specific QI programme successfully embedded evidence-based Golden Hour practices in a high-volume, resource-constrained maternity unit. Combining education, equipment, clinical support and continuous data feedback achieved sustained change in practice and a trend towards improved survival.

## Problem

Sally Mugabe Central Hospital (SMCH) hosts Zimbabwe’s busiest tertiary referral maternity unit with approximately 1,800 births per month. Its 13-bedded labour ward faces challenges including running overcapacity, staffing pressures and equipment and medication availability. The caesarean section rate sits between 20-25% with two functional obstetric theatres. The stillbirth rate averages at 50.9 per 1000 births, with 82% after 28 weeks gestation (SMCH, unpublished data 2026), above the country’s 2030 Every Newborn Action Plan of 12 or fewer per 1000 births^1^.

The World Health Organization recommends that all newborns not requiring immediate resuscitation should receive delayed cord clamping (DCC) for 1-3 minutes, early skin to skin care (ESSC) and initiation of breastfeeding within the first hour of life^2^. These interventions have a strong evidence base and particularly benefit low birth weight (<2500 grams, LBW) or premature (<37 weeks gestation) newborns^3^ but implementation is inconsistent across Sub-Saharan Africa (SSA). A review of newborn care practices in 14 hospitals in the African Neonatal Network (including Zimbabwe) in 2025 reported DCC in approximately 47.4% of newborns and 31.1% of newborns <32 weeks gestation^4^. In Zimbabwe, DCC and ESSC rates are not captured as national facility-level indicators and prior to this project there was no documentation of DCC or ESSC practice at SMCH.

Our project included a scoping phase utilising complementary sources of information to establish baseline practice. This included: existing knowledge of departmental workflow from the project team who were embedded within the department, direct observation of clinical practice across several shifts in theatre and on labour ward, using a structured checklist to record DCC and ESSC practice alongside informal observation of resuscitation practice. In addition, participatory discussions with clinical staff were held at the beginning of each training session to map current practice and barriers to change.

Significant variability in practice was identified; midwives reported they would occasionally carry out DCC or ESSC in well term newborns but this was not standard in a theatre setting or in LBW babies. Barriers included lack of time, concerns that small babies would get cold, staffing pressures, equipment shortages (eg. no clocks) and lack of recognition of the importance of these interventions. Routine practice on labour ward was to cut the umbilical cord immediately and bring the baby to the resuscitaire to be dried and weighed. The mother would usually be moved within ten minutes of delivering to a communal wooden bench. Babies would wait in the “Baby Room” until dressed, which could be between 30 minutes to several hours later depending on staff availability. Babies who required review by the paediatric team would be returned to their mothers once this had been completed.

Neonatal resuscitation was initiated by midwives with support from doctors called after resuscitation commenced. Resuscitation equipment was inconsistently available and there was no fully working resuscitaire or suction. The highest level of respiratory support was nasal cannula oxygen, supplied to a maximum of two babies at a time. Babies requiring transfer to the neonatal unit would be carried up the stairs by midwives with no transport incubator available.

SMCH’s attached 100 cot neonatal unit accepts babies born from within the hospital and nationwide for specialist care and surgical expertise and admits approximately 450-550 babies each month. The most common reasons for admission include prematurity, LBW, neonatal encephalopathy and sepsis. A 2018 study estimated a 55% mortality rate for very LBW newborns in the unit^5^ and a recent analysis of neonatal outcomes at SMCH (2021-2025) showed that pre-term babies admitted have 4-6 times higher odds of death than their term counterparts (Neotree, unpublished data, 2025).

The aims of this project were:

- To implement a package of interventions to improve the management of newborns in the first hour of life.
- To improve the rates of DCC and ESSC in eligible babies to 80% over a 12 month period.
- To maintain the proportion of babies admitted to the neonatal unit who were normothermic (36.5-37.5 degree celsius) at over 80%.
- To evaluate the impact of implementing the Golden Hour intervention package on the outcomes of a subset of babies admitted to the neonatal unit.

## Background

Globally in 2024, an estimated 4.9 million children died before the age of 5 years. Almost half of these were neonatal deaths in the first 28 days of life of which 50% occurred in SSA^6^. A 2026 analysis of the trajectory of neonatal mortality rates worldwide demonstrated a 54% slowdown of reduction rate in 2015-2024, with Southern Africa representing one of the slowest rates of progress globally ^7^. Zimbabwe, in this region, saw a rise in neonatal mortality rate to 36 per 1000 live births despite a SDG target to reduce neonatal mortality to 12 per 1000 live births by 2030.^8^ Low-cost, evidence-based interventions, when scaled up, have the potential to reduce up to 55% of neonatal deaths^9^. Despite recognition of the importance of such interventions in national guidelines such as the Zimbabwe Every Newborn Action Plan, implementation remains inconsistent.

DCC for 60-120 seconds is advocated by governing bodies to improve infant health, nutrition and outcomes.^2,10^ The most significant benefits are seen in preterm babies, including reduced rates of necrotising enterocolitis, hypotension, anaemia and an overall reduction in mortality of 30%^10-18^. ESSC is associated with improved temperature, oxygenation and heart rate stability^19,20^, improved breastfeeding rates, reduced neonatal sepsis and lower mortality in LBW babies^21-24^. A systematic review in 2025 mapped QI projects focusing on the first hour of life (“The Golden hour”) in preterm babies. The majority of studies published were from high income settings but showed that protocol adherence improved with QI projects with subsequent improvements in neonatal outcomes^25^.

## Measurement

This was a prospective QI project using iterative PDSA cycles. We used routinely collected clinical data to evaluate implementation and conducted a secondary observational analysis of neonatal mortality among eligible neonatal admissions.

Prior to our project the provision of DCC or ESSC at SMCH was not documented. A snapshot baseline observational audit of 23 deliveries in October 2024 found that 74% of babies were eligible for DCC and ESSC and these interventions were carried out in 41% of eligible deliveries. DCC was not timed and no LBW babies received DCC or ESSC.

We captured delivery information on all babies born at SMCH over 12 months (1st of November 2024 - 31st October 2025) using Neotree. Neotree is a digital quality improvement platform which has been embedded within neonatal care at SMCH since 2019^26-28^. The user-facing component is a tablet-based Android application which is used by healthcare providers to admit and discharge babies to and from the neonatal unit and provides clinical decision support, access to context specific guidelines and real time data capture. In addition, and to support the development of a perinatal version of Neotree, a research nurse also collects retrospective maternal and neonatal characteristics and outcome data for every delivery. We added questions to track DCC and ESSC progress into this retrospective data capture script for Neotree.

Our measure of intervention uptake was the number of eligible babies who had DCC or ESSC recorded. We defined DCC as clamping of the umbilical cord between 1-3 minutes after delivery and ESSC as intentional skin to skin contact with the mother occurring in the first hour of life for any duration. We determined babies as eligible for the interventions if they did not require immediate resuscitation. Quantitative data were supplemented by at least weekly clinical observations on labour ward and discussions with clinical staff during training sessions. Changes to newborn resuscitation practices were not formally measured.

In addition to tracking intervention uptake for all babies, we explored the impact of our project interventions on neonatal outcomes and temperature on admission for a smaller subset of babies admitted to the neonatal unit. We fitted a binomial multivariable logistic regression model on eligible infants admitted to the neonatal unit (born within SMCH, 1 min Apgar above 6) to analyse the effect of DCC and ESSC on neonatal mortality and adjusted this model for birth-weight and sex. The statistical analysis methods for this mortality association logistic regression are detailed fully in Supplementary materials.

## Design

Led by a neonatal consultant (MM) working at SMCH, our project team included a research nurse (PN), paediatric doctor (RP) and neonatal nurse (RS). Logistical, technical, statistical and data support, implementation guidance and academic oversight was provided by the wider Neotree project team in Zimbabwe and University College London (UCL) and Imperial College Neotree academic teams in the UK.

Given the clinical and contextual expertise within the project team we were aware of the challenges in providing care in the first hour of life at SMCH (as detailed in the ‘Problem’ section above). We reviewed the journey from a mother’s arrival to baby’s birth, utilising QI methodology to map out factors which impacted care allowing us to develop highly context-specific interventions. We met with the obstetric and neonatal leadership teams, ward matrons and other clinical staff at SMCH to share our plans for the project and incorporated their comments and feedback into our interventions.

### Patient involvement

Mothers and their families were not directly involved in setting the quality improvement focus or the outcome measure however we included informal input from inpatient mothers in designing practical strategies for improving ESSC and thermal care on labour ward and theatres. We also gained the opinions of a community engagement group on educational posters displayed to describe the benefits of ESSC.

### Ethics

We gained ethical approval from SMCH ethics committee (Ref:SMCHE180924/94) and data on neonatal outcomes were extracted in anonymised form from Neotree with existing ethical approval from Biomedical Research and Training Institute (AP171/2022), the Medical Research Council of Zimbabwe, (MRCZ/A/2570), the Joint Research Ethics Committee review board (JREC/327/19) and the UCL Research Ethics Committee (26279/001).

Our interventions included:

- **Training**. An educational package was developed as a series of interactive workshops covering the scientific basis, practical aspects and benefits of DCC, ESSC, newborn resuscitation, transfer to the neonatal unit and thermal care. We delivered two sessions every two months to capture the large number of maternity staff. An adapted version with a focus on newborn resuscitation was delivered to doctors working on NICU.
- **Equipment**. We carried out a needs assessment and provided equipment to support DCC, ESSC, thermal care and newborn resuscitation including: heaters, clocks, timers, plastic bags, hats, blankets, chairs, bag valve masks and nasal cannula.
- **Feedback and progress tracking**. We held regular meetings to share progress with staff. This included a mixture of informal updates, posters with ‘what is going well’ and ‘things to work on’ messaging around labour ward and theatres, formal presentations at departmental meetings and posts on the labour ward staff WhatsApp group. To encourage and recognise staff we timed these updates with the provision of a food-based hamper (containing $5 of pasta, flour, sugar) every quarter. This was based on previous successful use of incentives in the Neotree implementation^28^ and following positive feedback after our first trial hamper at Christmas.
- **Clinical Support**. Clinical support was provided by members of the project team (pediatrician, neonatologist or paediatric nurse) to deliver hands-on education and peer support, build ongoing relationships with health care workers and gain greater understanding of the practical challenges in delivering care in the first hour of life. For the first 1-6 months of the project this involved at least twice weekly presence in labour ward or theatres, spaced to at least once weekly by months 6 -12.

Anticipated challenges at the planning stage were the rotation and number of clinical staff, high midwife/patient ratios, sustaining enthusiasm and motivation for the project and logistical barriers such as equipment availability and staff shortages. To ensure sustainability we embedded the project within existing record keeping systems, trained permanent clinical staff at SMCH, provided equipment and shared teaching materials with the midwifery school.

## Strategy

### PDSA 1 – Documentation (Table 1)

At delivery, midwives document infant birth details on paper notes without prompts to record DCC/ESSC status. During our first workshops midwives decided the best place to record this information and developed a system of documenting this in the formal delivery log to enable retrospective review by our research nurse. In the first month we noticed the DCC/ESSC status was only recorded in 28% of births. At subsequent training, feedback meetings and during clinical support sessions we emphasised the importance of this information and provided the option to add a reason for not performing these interventions. By month two a yes/no status was recorded for 70.7% of babies and then maintained between 78.8-92.4% from month four onwards. Of the 22,036 live births over the 12 months, 18,854 (85.6%) were eligible for the interventions. In the 3,182 ineligible infants, the majority appropriately did not receive either intervention (2,315, 72.8%).

**Table 1.** Documentation of DCC and ESSC status in all live births over each month of the project.

| Month | Live births | Eligible* | DCC Yes | DCC No | DCC Unknown | DCC status documented (% of live births) | ESSC Yes | ESSC No | ESSC Unknown | ESSC status documented (% of live births) |
|---|---|---|---|---|---|---|---|---|---|---|
| Nov-24 | 1,735 | 834 | 402 | 83 | 1250 | 28.00% | 378 | 104 | 1253 | 27.80% |
| Dec-24 | 1,723 | 1,519 | 1,088 | 131 | 504 | 70.70% | 1,077 | 132 | 514 | 70.20% |
| Jan-25 | 1,976 | 1,779 | 1,471 | 125 | 380 | 80.80% | 1,410 | 181 | 385 | 80.50% |
| Feb-25 | 1,728 | 1,523 | 1,187 | 174 | 367 | 78.80% | 1,178 | 182 | 368 | 78.70% |
| Mar-25 | 1,893 | 1,665 | 1,465 | 142 | 286 | 84.90% | 1,448 | 157 | 288 | 84.80% |
| Apr-25 | 1,841 | 1,628 | 1,520 | 90 | 231 | 87.50% | 1,457 | 150 | 234 | 87.30% |
| May-25 | 1,906 | 1,712 | 1,470 | 121 | 315 | 83.50% | 1,408 | 180 | 318 | 83.30% |
| Jun-25 | 1,867 | 1,622 | 1,569 | 130 | 168 | 91.00% | 1,504 | 193 | 170 | 90.90% |
| Jul-25 | 1,931 | 1,747 | 1,565 | 135 | 231 | 88.00% | 1,445 | 255 | 231 | 88.00% |
| Aug-25 | 1,843 | 1,651 | 1,517 | 161 | 165 | 91.00% | 1,341 | 336 | 166 | 91.00% |
| Sep-25 | 1,752 | 1,549 | 1,344 | 161 | 247 | 85.90% | 1,162 | 343 | 247 | 85.90% |
| Oct-25 | 1,841 | 1,625 | 1,557 | 144 | 140 | 92.40% | 1,366 | 335 | 140 | 92.40% |
| **All** | **22,036** | **18,854** | **16,155** | **1,597** | **4,284** | **80.60%** | **15,174** | **2,548** | **4,314** | **80.40%** |

*\*Eligible = “Did baby require immediate resuscitation? = No”*

### PDSA 2 – DCC and ESSC for LBW babies

There was reluctance to carry out DCC and ESSC in LBW babies as the midwives felt they were particularly fragile and did not want to delay transfer to the neonatal unit. We highlighted the benefit of the interventions and how to perform them safely in this population and saw an improvement in the uptake of ESSC from 31% (month 1) to 54-77% (months 3-12) and DCC from 33% (month 1) to 72-86% (months 5-12). (Figure 1 and Supplementary tables S1a and S1b.)

**Figure 1.**
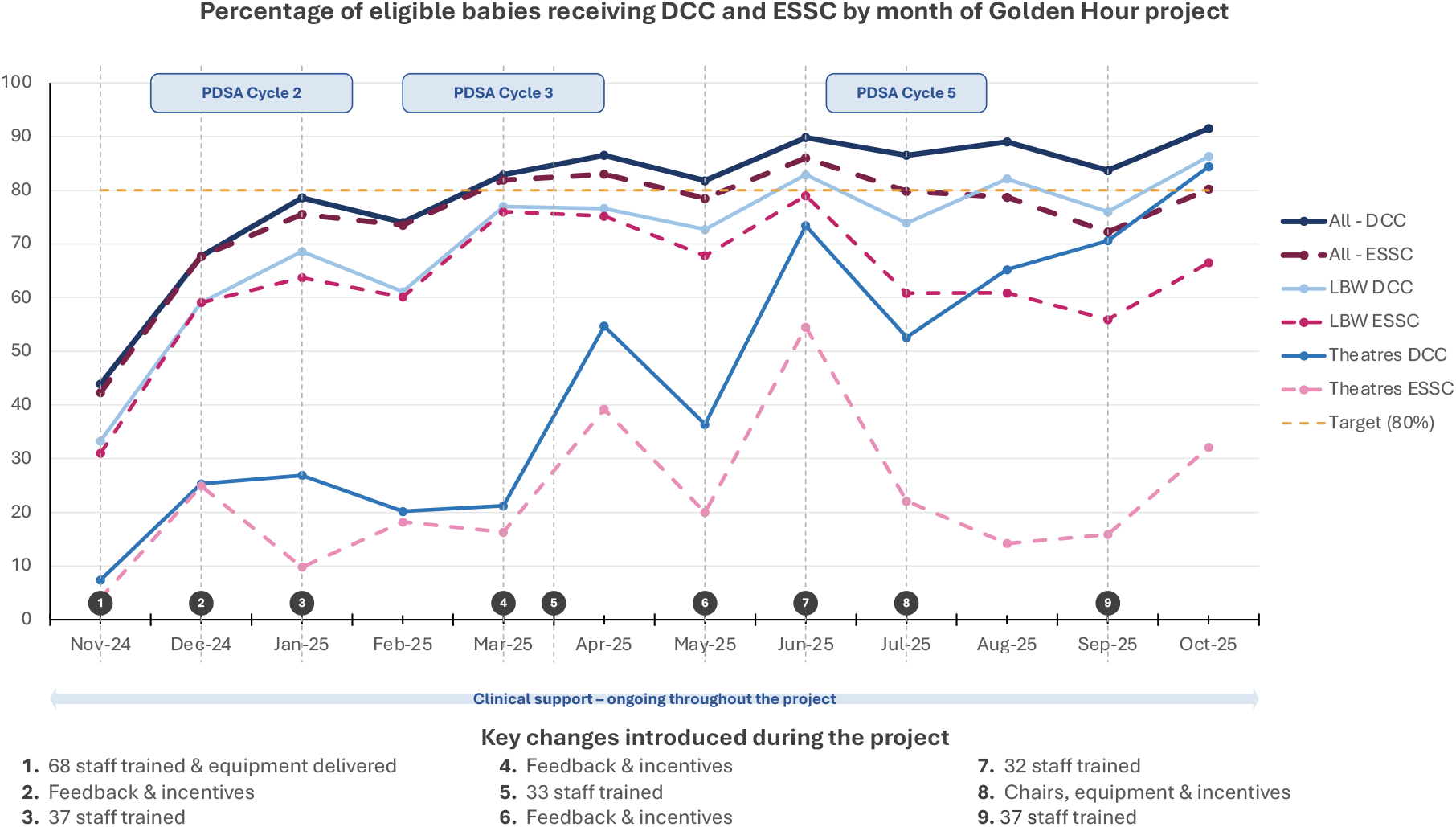
Percentage of eligible infants receiving DCC and ESSC (all infants compared to low birth weight and those born in theatres) alongside key QI interventions during the 12 month implementation period.

### PDSA 3 – DCC and ESSC for babies born in theatres

We noted a discrepancy in implementation between babies born in theatre compared to labour ward, with only a quarter of babies in theatre receiving DCC and ESSC compared to over 80% on labour ward. Key themes for the low implementation in theatres included: logistical challenges maintaining a sterile field, high patient turn-over limiting time for DCC and ESSC, equipment challenges with a lack of beds to facilitate safe ESSC, many unwell mothers and a reticence to challenge the clinician performing the caesarean section. In response we included a demonstration of DCC and ESSC within a sterile field in our training sessions, held training for obstetric doctors, attended theatre to support and demonstrate these processes and provided equipment including hats, clocks and heaters to theatre areas. DCC rates in theatres improved to 52% by month 6 and to 84% by month 12. ESSC rates rose to a peak of 53% (June 25) but were harder to sustain. (Figure 1 and Supplementary table S2a and S2b.)

### PDSA 4 – Thermal care

Month seven of our project fell during winter, increasing the urgency to improve thermal care for neonates. We were not able to track the temperature of every baby delivered, but we reviewed the temperature of babies born within SMCH who were subsequently admitted to the neonatal unit. The percentage of babies normothermic on admission assessment (completed on labour ward) fell from 85-92% in warmer Nov-Feb to 72% in Apr 2025. We observed that babies were not being fully dried or given a hat until they were formally dressed. The labour ward had no heating and broken windows resulted in cold temperatures. We provided three ‘Warmilu’ blankets adapted to utilise hot water bottles to ensure the blanket could be kept reliably warm, installed radiant heaters in labour ward, set up a fundraising initiative to source blankets and introduced a “Hat with your (delivery) pack” messaging for labour ward and “Hat with the notes” system for theatres. The percentage of babies with a normal temperature improved and remained between 81-83% for the rest of the colder months (May-July). (Supplementary - Figure S1)

### PDSA 5 – Extending ESSC and reducing separation of mother and baby

Despite ESSC rates increasing this was often carried out for only a few minutes before babies were separated to wait to be dressed, for doctor review or transfer to postnatal ward. This impacted thermal care, initiation of breastfeeding and limited the benefits of ESSC. We introduced a baton on-call phone to enable midwives to reach doctors more quickly for review and highlighted the benefits of keeping babies with their mothers unless very unwell. We obtained a donation of chairs designed to accommodate a reclining mother carrying out skin to skin care and designed posters sharing the benefits of ESSC and breastfeeding in Shona and English. (Figure 1)

## Results

### Training and uptake of interventions

During the 12 month project we delivered 12 training sessions to 207 clinical staff and 87 midwifery students in addition to at least weekly bedside training and mentoring sessions. A total of 15,292 babies (81.1% of eligible) were documented as having received DCC and 14,351 (76.1%) as receiving ESSC of any duration. The characteristics of eligible infants and their DCC/ESSC status is shown in Supplementary Table S3.

Figure 1 demonstrates the uptake of DCC and ESSC in all babies by month compared to uptake in those born in theatre and LBW babies, alongside relevant QI interventions and cycles. Our documented implementation of DCC and ESSC in all eligible babies increased from 44% to 68% in the first two months of the project to over 80% in March 2025 before settling between 82-91% DCC and 72-85% ESSC for the remaining seven months of the project, with DCC implementation consistently higher than ESSC.

### Impact of interventions on neonatal mortality

Between the 1st November 2024 to 31st October 2025, there were 22,036 live births, of which 4,860 infants were admitted to the neonatal unit. Excluding 1,770 infants with a 1 minute Apgar score <7 left 3,090 eligible infants. A documented DCC/ESSC status was available for 1,794 (58.1%) of these infants of whom 1,654 infants had a linked discharge record allowing mortality analysis. Among these 1,794 infants, 920 (51.3%) received neither intervention, 426 (23.7%) DCC only, 61 (3.4%) ESSC only, and 387 (21.6%) both. Birth-weight and gestational-age distributions were broadly similar across groups, with a slightly higher proportion of term infants and infants delivered vaginally in the both-interventions group. (Supplementary Table S4.)

Of the 1654 infants with a linked discharge record, death before discharge occurred in 108 infants (6.5%, 95% CI 5.4–7.8) overall. Crude mortality was 6.6% (95% CI 5.1–8.5) in the neither group, 7.3% (5.0–10.1) in the DCC-only group, 10.3% (4.8–20.8) in the ESSC-only group and 5.0% (3.2–7.7) in the both-interventions group. In logistic-regression models, receiving both interventions was associated with lower odds of death before discharge but the association did not reach statistical significance. Compared with the neither group, the odds ratio for both interventions was 0.74 (95% CI 0.43–1.27) unadjusted and 0.93 (0.52–1.66) after adjustment for birth weight and sex, and 0.81 (0.36–1.80) in the <2500g subgroup. (Table 2, Supplementary Figure S2) A sensitivity analysis restricted to infants of 1500–2499g was directionally consistent but confidence intervals crossed the null. (Supplementary Table S5).

**Table 2.**
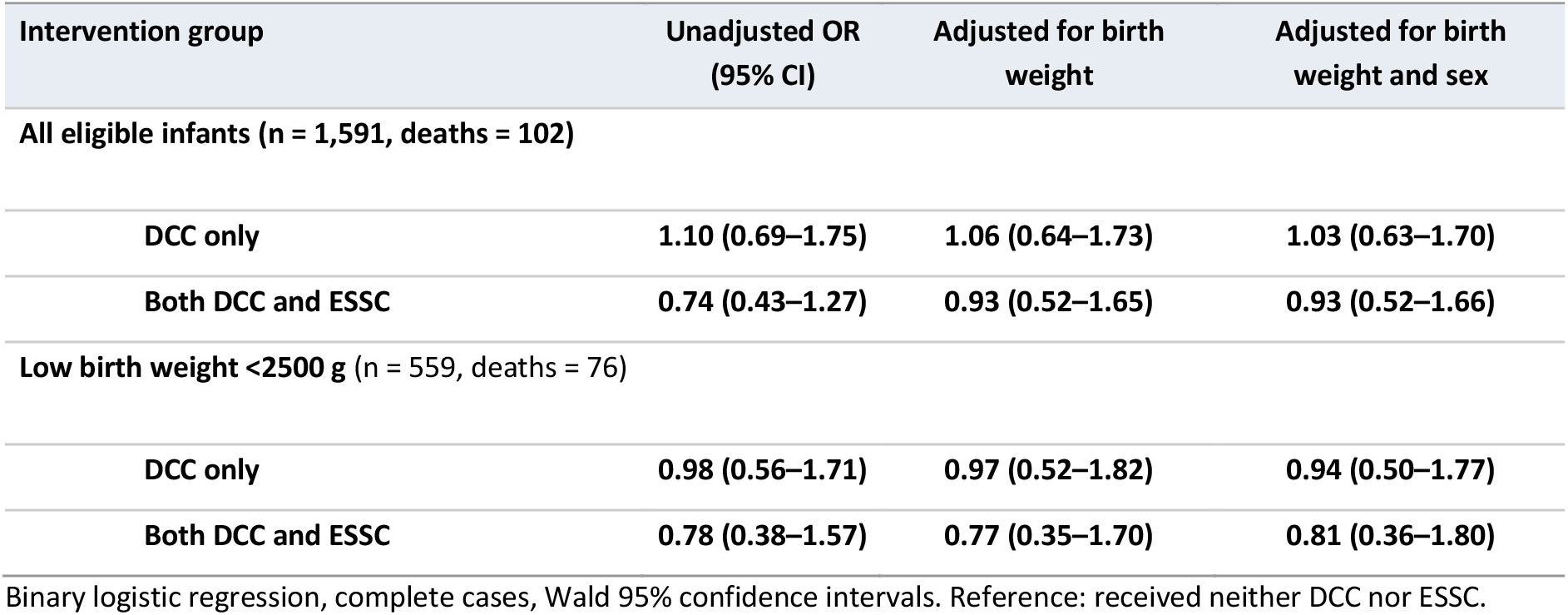
Odds ratios for death before discharge by intervention group, all eligible infants with a matched discharge outcome and low-birth-weight subgroup (excluding ESSC-only group, n = 58)

## Lessons and limitations

This project resulted in an increased and sustained implementation of DCC and ESSC in a busy central hospital in Zimbabwe and provided training and equipment to support newborn resuscitation and thermal care.

We recognised that implementation was limited by multiple factors, not only by lack of awareness, and there was significant complexity in overcoming the implementation gap to deliver so-called simple interventions. We had to employ various strategies to ensure that our interventions were highly context specific and work collaboratively with clinical staff to understand the multi-factorial barriers to providing optimum care. For example, enabling ESSC beyond the first minute of life required creative and ‘next-best practice’ solutions with the acceptance of a short break in skin-to-skin care while a mother moved to the chair area after it became apparent that the acuity on labour ward rarely allowed a mother to stay in the delivery bed. In some cases social expectations resulted in unexpected barriers, requiring sensitive untangling to enable a change of practice. For example, the expectation that the hat and towels brought by the mother should only be used once the baby was already clean and dressed led to babies spending several hours in wet linen with no hat before a midwife had time to dress them.

Another learning point was that the provision of equipment to a resource and infrastructure constrained setting was not enough in isolation to improve clinical care. Resuscitation equipment was often misplaced and maintenance of even low-cost equipment such as batteries in clocks proved challenging. The multipronged approach of our project providing equipment, educational workshops, informal discussion and best practice demonstrations enabled us to overcome some, but not all, of these challenges.

An important factor in the success of our project was the engagement, involvement and support from obstetric and neonatal consultants, hospital matrons, theatre staff, midwifery school lecturers and midwives. Key to this was our project team composition which included a departmental lead neonatologist and neonatal nurse who were well respected within the hospital and our clinical volunteers spent a minimum of six months on site to become embedded within the team. Maintaining a clinical presence on labour ward also provided invaluable teaching opportunities, insight into challenges faced by clinical staff and built trust that there was investment in the project. We also found that informal feedback methods, such as updates on the labour ward WhatsApp group and provision of food-based hampers to recognise staff effort in a demanding, resource-constrained system, were well received and helped sustain engagement.

Embedding our data collection within an existing tool (Neotree) was a strength of our project as it enabled us to track and respond to real-time trends in our implementation without adding a significant data collection burden. This also increased sustainability by enabling ongoing monitoring of indicators after the formal project ended. Limitations include documenting reported practice which may not always reflect true actions. We were reassured that our reported implementation rates correlated with actual practice by our clinical observations, the fact that less babies received interventions during busy periods (suggesting a true recording of actions rather than a box ticking exercise) and the fact that babies with a low Apgar score had DCC and ESSC appropriately recorded as not carried out. Some of our noted increase in uptake may also represent an improvement in documentation.

Our newborn resuscitation training was not formally evaluated as part of this project, and thermal care was only assessed among the subset of babies admitted to the neonatal unit rather than all deliveries. The intervention initially achieved a partial recovery from a seasonal dip in neonatal thermal care, although this was measured only among babies admitted to the neonatal unit and was not sustained. We were reluctant to increase data collection burden for an already stretched workforce and so accepted that some potentially valuable data points such as temperature on every baby were not possible to collect.

The primary focus of this project was QI and therefore the study was not powered to detect a statistically significant difference in neonatal outcome in the smaller subset of infants admitted to the neonatal unit. Our population size was further limited by more incomplete documentation of DCC/ESSC status in the admission process compared to that in the delivery register. Subset analysis of this admitted group found a non-significant lower crude mortality among infants receiving both DCC and ESSC.

## Conclusion

A multifaceted QI approach, combining staff training, leadership engagement, regular staff consultation, equipment provision, clinical support, and real-time feedback, improved uptake of DCC and ESSC in a busy, resource-constrained tertiary maternity unit in Zimbabwe. Improvements were sustained over 12 months and were mirrored in groups where implementation was particularly challenging, including LBW babies and those born in theatre.

Our findings illustrate that implementing evidence-based newborn interventions requires more than equipment or education alone. Understanding and addressing contextual barriers, with staff engaged throughout and data used to identify gaps as they arise, is key to achieving sustained change in newborn care.

Next steps include continuing to consolidate the implementation of DCC and ESSC, alongside an intervention to improve thermal care during transfer from the labour ward to the neonatal unit. We also plan to expand the Neotree clinical decision support and data collection tool to include the perinatal period rather than the neonatal period alone, which will help identify infants at greatest risk to enable targeted future interventions to improve neonatal morbidity and mortality.

## Data Availability

All data produced in the present study are available upon reasonable request to the authors and following subsequent authorisation.

https://github.com/neotree/golden-hour

## Acknowledgments

We are hugely grateful to the staff and families at SMCH, whose dedication and daily care this project reflects. With thanks also to the Neotree charity and whole Neotree project team.

## Funding statement

This material has been funded by UK International Development from the UK government; NURTURE Zimbabwe: Neotree Technology Enabled Newborn Care and Research Project (Grant Number: 300816-402). MH is supported by a NIHR Global Health Professorship (NIHR302422). FF is supported by a Wellcome Trust Early Career Award (227076/Z/23/Z). The findings and views expressed are those of the authors and do not necessarily reflect the UK government official policies or those of NIHR or the Wellcome Trust.

## Competing Interest Statement

FF and MH are trustees of Neotree, a UK registered charity that provides technology, software information, education and support to healthcare workers and medical practitioners throughout England and Wales, Malawi and Zimbabwe (charity number: 1186748). MH is additionally a co-founder of Neotree. All other authors declare no competing interests.

## Contributorship Statement

Project concept and design: MM, RP, FF, SC and HG. Project clinical/technical team in SMCH: MM, RP, PN, Kma and RS. Monthly project data review, analysis and reporting: RP, MM, PN, KMe, CM, KMa, MH, SC, RS, HG and FF. End of project data analysis: DL, RP and MM. Preliminary draft: RP, MM, DL, MH and FF. Final draft: all authors.

FF and MM are joint senior authors.

## References

1. World Health Organization, UNICEF. Every newborn: an action plan to end preventable deaths. Geneva: World Health Organization 2014. https://www.who.int/publications-detail-redirect/9789241507448 (accessed 29/09/2026)

2. World Health Organization. Guideline: delayed umbilical cord clamping for improved maternal and infant health and nutrition outcomes. Geneva: World Health Organization 2014. https://www.who.int/publications-detail-redirect/9789241508209 (accessed 01/06/2026)

3. Seidler AL, Aberoumand M, Hunter KE, et al. Deferred cord clamping, cord milking, and immediate cord clamping at preterm birth: a systematic review and individual participant data meta-analysis. Lancet 2023;402:2209–22. doi: 10.1016/S0140-6736(23)02468-6

4. Stevenson AG, Ehret DE, Abayneh M, et al. Characteristics, interventions, and status of infants discharged from 14 hospitals in the African Neonatal Network, 2024. J Afr Neonatol 2025;3:68–77. doi: 10.4314/jan.v3i3.2

5. Zvenyika-Hlatywayo LT, Chimhuya S, Gumbo FZ, et al. Risk factors for mortality in low birth weight infants at Harare Hospital (Maternity Unit), Zimbabwe. Clin Pediatr Res 2018;2:33–41.

6. Sharrow D, Hug L, You D, et al. Global, regional, and national trends in under-5 mortality between 1990 and 2019 with scenario-based projections until 2030: a systematic analysis by the UN Inter-agency Group for Child Mortality Estimation. Lancet Glob Health 2022;10:e195–206. doi: 10.1016/S2214-109X(21)00515-5

7. Sharrow D, Hug L, Liu Y, et al. Global, regional, and national levels and trends in under 5, infant, and neonatal mortality during 1990-2024 with scenario based projections to 2030: modelling study. BMJ 2026;393:e088684. doi: 10.1136/bmj-2025-088684

8. Zimbabwe National Statistics Agency (ZIMSTAT), ICF. Zimbabwe Demographic and Health Survey 2023–24: Key indicators report. Harare, Zimbabwe, and Rockville, Maryland, USA: ZIMSTAT and ICF 2024. https://dhsprogram.com/publications/publication-pr160-preliminary-reports-key-indicators-reports.cfm (accessed 29/9/2026)

9. Darmstadt GL, Bhutta ZA, Cousens S, et al. Evidence-based, cost-effective interventions: how many newborn babies can we save? Lancet 2005;365:977–88. doi:10.1016/S0140-6736(05)71088-6

10. McAdams RM, Lakshminrusimha S. Management of placental transfusion to neonates after delivery. Obstet Gynecol 2022;139:121–37. doi: 10.1097/AOG.0000000000004625

11. Fogarty M, Osborn DA, Askie L, et al. Delayed vs early umbilical cord clamping for preterm infants: a systematic review and meta-analysis. Am J Obstet Gynecol 2018;218:1–18. doi: 10.1016/j.ajog.2017.10.231

12. Jasani B, Torgalkar R, Ye XY, et al. Association of umbilical cord management strategies with outcomes of preterm infants: a systematic review and network meta-analysis. JAMA Pediatr 2021;175:e210102. doi: 10.1001/jamapediatrics.2021.0102

13. Persad E, Sibrecht G, Ringsten M, et al. Interventions to minimize blood loss in very preterm infants: a systematic review and meta-analysis. PLoS One 2021;16:e0246353. doi: 10.1371/journal.pone.0246353

14. Rabe H, Gyte GM, Díaz-Rossello JL, et al. Effect of timing of umbilical cord clamping and other strategies to influence placental transfusion at preterm birth on maternal and infant outcomes. Cochrane Database Syst Rev 2019;9:CD003248. doi: 10.1002/14651858.CD003248.pub4

15. Georgieff MK. Long-term brain and behavioral consequences of early iron deficiency. Nutr Rev 2011;69 Suppl 1:S43–8. doi: 10.1111/j.1753-4887.2011.00432.x

16. Gupta R, Ramji S. Effect of delayed cord clamping on iron stores in infants born to anemic mothers: a randomized controlled trial. Indian Pediatr 2002;39:130–5. PMID: 11867842

17. Chaparro CM, Neufeld LM, Tena Alavez G, et al. Effect of timing of umbilical cord clamping on iron status in Mexican infants: a randomised controlled trial. Lancet 2006;367:1997–2004. doi: 10.1016/S0140-6736(06)68889-2

18. KC A, Rana N, Målqvist M, et al. Effects of delayed umbilical cord clamping vs early clamping on anemia in infants at 8 and 12 months: a randomized clinical trial. JAMA Pediatr 2017;171:264–70. doi: 10.1001/jamapediatrics.2016.3971

19. Gupta N, Deierl A, Hills E, et al. Systematic review confirmed the benefits of early skin-to-skin contact but highlighted lack of studies on very and extremely preterm infants. Acta Paediatr 2021;110:2310–5. doi: 10.1111/apa.15913

20. Chi Luong K, Long Nguyen T, Huynh Thi DH, et al. Newly born low birthweight infants stabilise better in skin-to-skin contact than when separated from their mothers: a randomised controlled trial. Acta Paediatr 2016;105:381–90. doi: 10.1111/apa.13164

21. WHO Immediate KMC Study Group. Immediate “kangaroo mother care” and survival of infants with low birth weight. N Engl J Med 2021;384:2028–38. doi: 10.1056/NEJMoa2026486

22. Moore ER, Bergman N, Anderson GC, et al. Early skin-to-skin contact for mothers and their healthy newborn infants. Cochrane Database Syst Rev 2016;11:CD003519. doi: 10.1002/14651858.CD003519.pub4

23. Hendricks-Muñoz KD, Xu J, Parikh HI, et al. Skin-to-skin care and the development of the preterm infant oral microbiome. Am J Perinatol 2015;32:1205–16. doi: 10.1055/s-0035-1552941

24. Mitha A, Marchand-Martin L, Zores C, et al. Association between skin-to-skin contact and cognition and behaviour at 5 years in children born at 24–31 weeks’ gestation: a secondary analysis of a national population-based prospective cohort study. eClinicalMedicine 2025;89:103528. doi: 10.1016/j.eclinm.2025.103528

25. Wever LMC, Nous V, Hogeveen M. Golden hour(s) quality improvement protocols and their effect on outcomes in preterm infants ≤32 weeks of gestation: a scoping review. BMC Pediatr 2026;26:145. doi: 10.1186/s12887-025-06464-0

26. Heys M, Kesler E, Sassoon Y, et al. Development and implementation experience of a learning healthcare system for facility based newborn care in low resource settings: the Neotree. Learn Health Syst 2023;7:e10310. doi: 10.1002/lrh2.10310

27. Khan N, Crehan C, Hull-Bailey T, et al. Software development process of Neotree: a data capture and decision support system to improve newborn healthcare in low-resource settings. Wellcome Open Res 2022;7:305. doi: 10.12688/wellcomeopenres.18423.1

28. Gannon H, Chimhuya S, Chimhini G, et al. Electronic application to improve management of infections in low-income neonatal units: pilot implementation of the NeoTree beta app in a public sector hospital in Zimbabwe. BMJ Open Qual 2021;10:e001043. doi: 10.1136/bmjoq-2020-001043

