## Supplementary material for "The Golden Hour – Implementing and evaluating a package of interventions to improve newborn care in the first hour of life in a public sector hospital in Zimbabwe"

**Statistical Analysis Methods for Subset of Infants Admitted to the Neonatal Unit**

The study population was all inborn neonates admitted to the SMCH neonatal unit between 1 November 2024 and 31 October 2025, excluding readmissions. To restrict the comparison to infants for whom delayed cord clamping (DCC) and early skin-to-skin care (ESSC) were not contraindicated, infants with a 1-minute Apgar score below 7 were excluded; resuscitation type was not used to define eligibility because it was incompletely recorded (present for 16.0% of the cohort). Eligible infants were grouped by the delivery-room interventions documented in the Neotree record as having received neither DCC nor ESSC (reference), DCC only, ESSC only, or both. The outcome was death before discharge (neonatal death), analysed in infants with a linked discharge record. Odds ratios were estimated by binary logistic regression (unadjusted first), then adjusted for birth weight, then adjusted for birth weight and sex, fitted on complete cases. Analyses were repeated in the low-birth-weight subgroup (<2500 g) and, as a sensitivity analysis, in infants weighing 1500–2499g. The ESSC-only group was small and was summarised as counts only. 95% confidence intervals for crude mortality proportions were calculated using the Wilson score method.

**Software and reproducibility**

All analyses were performed in R version 4.6 (tidyverse 2.0.0) using the tidyverse, lubridate, scales, broom, officer and flextable packages. Each analysis is a self-contained, reproducible script (available at https://github.com/neotree/golden-hour). Statistical significance was assessed at a two-sided alpha of 0.05; because the study is a quality-improvement evaluation rather than confirmatory hypothesis testing, no adjustment for multiple comparisons was made.

### Additional Figures and Tables

**Table S1a.** Delayed cord clamping among eligible low birth weight infants (<2500g), by month (1 November 2024 – 31 October 2025)

| Month | Eligible low birth weight infants, n | Received, n (% of eligible) | Not received, n (%) | Unknown / not recorded, n (%) |
|---|---|---|---|---|
| Nov 2024 | 84 | 28 (33.3%) | 2 (2.4%) | 54 (64.3%) |
| Dec 2024 | 149 | 88 (59.1%) | 5 (3.4%) | 56 (37.6%) |
| Jan 2025 | 242 | 165 (68.2%) | 8 (3.3%) | 69 (28.5%) |
| Feb 2025 | 200 | 121 (60.5%) | 9 (4.5%) | 70 (35.0%) |
| Mar 2025 | 207 | 159 (76.8%) | 4 (1.9%) | 44 (21.3%) |
| Apr 2025 | 205 | 154 (75.1%) | 2 (1.0%) | 49 (23.9%) |
| May 2025 | 219 | 158 (72.1%) | 3 (1.4%) | 58 (26.5%) |
| Jun 2025 | 191 | 155 (81.2%) | 6 (3.1%) | 30 (15.7%) |
| Jul 2025 | 213 | 156 (73.2%) | 5 (2.3%) | 52 (24.4%) |
| Aug 2025 | 174 | 142 (81.6%) | 7 (4.0%) | 25 (14.4%) |
| Sep 2025 | 171 | 128 (74.9%) | 5 (2.9%) | 38 (22.2%) |
| Oct 2025 | 190 | 163 (85.8%) | 2 (1.1%) | 25 (13.2%) |
| **TOTAL** | **2,245** | **1,617 (72.0%)** | **58 (2.6%)** | **570 (25.4%)** |

**Table S1b.** Early skin to skin care among eligible low birth weight infants (<2500g), by month (1 November 2024 – 31 October 2025).

| Month | Eligible low birth weight infants, n | Received, n (% of eligible) | Not received, n (%) | Unknown / not recorded, n (%) |
|---|---|---|---|---|
| Nov 2024 | 84 | 26 (31.0%) | 4 (4.8%) | 54 (64.3%) |
| Dec 2024 | 149 | 88 (59.1%) | 5 (3.4%) | 56 (37.6%) |
| Jan 2025 | 242 | 153 (63.2%) | 18 (7.4%) | 71 (29.3%) |
| Feb 2025 | 200 | 119 (59.5%) | 11 (5.5%) | 70 (35.0%) |
| Mar 2025 | 207 | 157 (75.8%) | 6 (2.9%) | 44 (21.3%) |
| Apr 2025 | 205 | 151 (73.7%) | 5 (2.4%) | 49 (23.9%) |
| May 2025 | 219 | 147 (67.1%) | 14 (6.4%) | 58 (26.5%) |
| Jun 2025 | 191 | 147 (77.0%) | 14 (7.3%) | 30 (15.7%) |
| Jul 2025 | 213 | 127 (59.6%) | 33 (15.5%) | 53 (24.9%) |
| Aug 2025 | 174 | 104 (59.8%) | 45 (25.9%) | 25 (14.4%) |
| Sep 2025 | 171 | 92 (53.8%) | 41 (24.0%) | 38 (22.2%) |
| Oct 2025 | 190 | 124 (65.3%) | 41 (21.6%) | 25 (13.2%) |
| **TOTAL** | **2,245** | **1435 (63.9%)** | **237 (10.6%)** | **573 (25.5%)** |

Eligible = "Did baby require immediate resuscitation? = No". Low birth weight = birthweight at discharge <2500g. "% of eligible" counts unknown and not-recorded status in the denominator; "% of documented" uses only records coded Y or N.

**Table S2a.** Delayed cord clamping among eligible caesarean live births, by month (1 November 2024 – 31 October 2025)

| Month | Eligible caesarean births, n | Received, n (% of eligible) | Not received, n (%) | Unknown / not recorded, n (%) |
|---|---|---|---|---|
| Nov 2024 | 149 | 11 (7.4%) | 3 (2.0%) | 135 (90.6%) |
| Dec 2024 | 265 | 67 (25.3%) | 0 (0.0%) | 198 (74.7%) |
| Jan 2025 | 303 | 81 (26.7%) | 5 (1.7%) | 217 (71.6%) |
| Feb 2025 | 282 | 51 (18.1%) | 26 (9.2%) | 205 (72.7%) |
| Mar 2025 | 317 | 64 (20.2%) | 3 (0.9%) | 250 (78.9%) |
| Apr 2025 | 310 | 161 (51.9%) | 0 (0.0%) | 149 (48.1%) |
| May 2025 | 348 | 120 (34.5%) | 1 (0.3%) | 227 (65.2%) |
| Jun 2025 | 309 | 224 (72.5%) | 2 (0.6%) | 83 (26.9%) |
| Jul 2025 | 371 | 193 (52.0%) | 1 (0.3%) | 177 (47.7%) |
| Aug 2025 | 334 | 216 (64.7%) | 2 (0.6%) | 116 (34.7%) |
| Sep 2025 | 326 | 229 (70.2%) | 0 (0.0%) | 97 (29.8%) |
| Oct 2025 | 336 | 281 (83.6%) | 1 (0.3%) | 54 (16.1%) |
| **TOTAL** | **3,650** | **1,698 (46.5%)** | **44 (1.2%)** | **1,908 (52.3%)** |

**Table S2b.** Early skin-to-skin care among eligible caesarean live births, by month (1 November 2024 – 31 October 2025)

| Month | Eligible caesarean births, n | Received, n (% of eligible) | Not received, n (%) | Unknown / not recorded, n (%) |
|---|---|---|---|---|
| Nov 2024 | 149 | 6 (4.0%) | 8 (5.4%) | 135 (90.6%) |
| Dec 2024 | 265 | 66 (24.9%) | 1 (0.4%) | 198 (74.7%) |
| Jan 2025 | 303 | 29 (9.6%) | 52 (17.2%) | 222 (73.3%) |
| Feb 2025 | 282 | 45 (16.0%) | 31 (11.0%) | 206 (73.0%) |
| Mar 2025 | 317 | 48 (15.1%) | 18 (5.7%) | 251 (79.2%) |
| Apr 2025 | 310 | 108 (34.8%) | 52 (16.8%) | 150 (48.4%) |
| May 2025 | 348 | 61 (17.5%) | 58 (16.7%) | 229 (65.8%) |
| Jun 2025 | 309 | 163 (52.8%) | 62 (20.1%) | 84 (27.2%) |
| Jul 2025 | 371 | 77 (20.8%) | 118 (31.8%) | 176 (47.4%) |
| Aug 2025 | 334 | 43 (12.9%) | 175 (52.4%) | 116 (34.7%) |
| Sep 2025 | 326 | 49 (15.0%) | 180 (55.2%) | 97 (29.8%) |
| Oct 2025 | 336 | 97 (28.9%) | 185 (55.1%) | 54 (16.1%) |
| **TOTAL** | **3,650** | **792 (21.7%)** | **940 (25.8%)** | **1,918 (52.5%)** |

*Eligible = did not require immediate resuscitation (resus = No), per the Table 1 eligibility definition. Caesarean = mode of delivery coded 4 or 5 (elective and emergency combined). "% of eligible" counts unknown and not-recorded status in the denominator.*

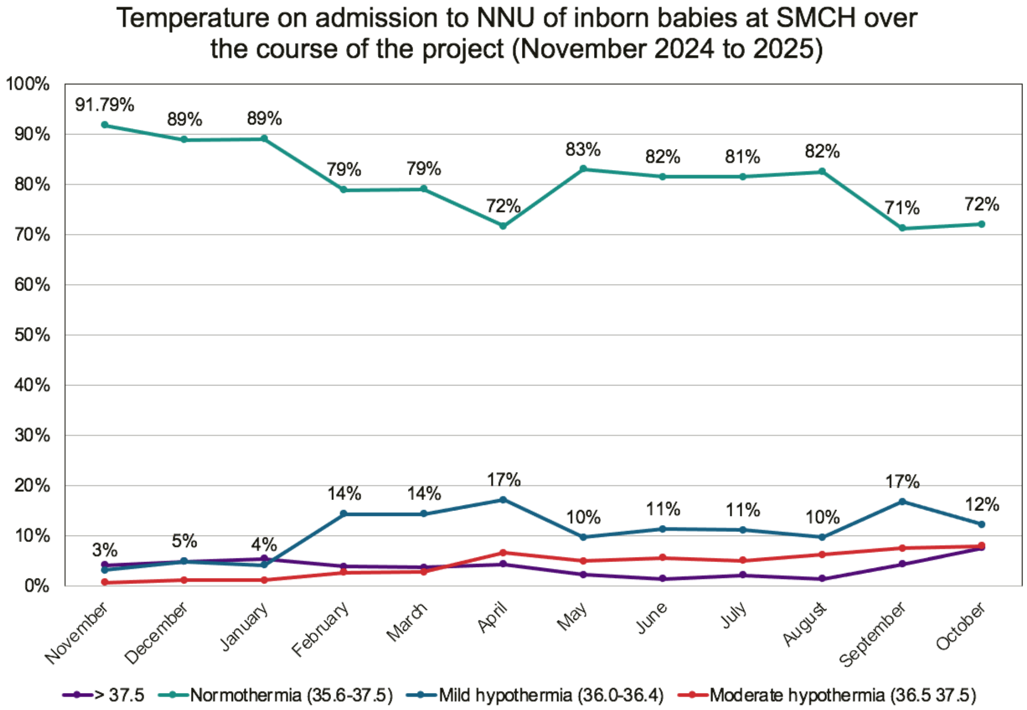


**Figure S1.** Percentage of inborn babies with a normal temperature on admission each month over the 12 month project period 1/11/2024 - 31/10/2025

**Table S3.** Uptake of delayed cord clamping (DCC) and early skin-to-skin care (ESSC) among eligible live births, by mode of delivery, birth weight and gestational age (SMCH, 1 November 2024 –31 October 2025)

| Characteristic | Eligible infants, n | Received DCC, n (%) | Received ESSC, n (%) | Received both DCC and ESSC, n (%) |
|---|---|---|---|---|
| Overall | 18,854 | 15,292 (81.1%) | 14,351 (76.1%) | 14,337 (76.0%) |
| Mode of delivery | | | | |
| Vaginal (incl. instrumental) | 15,204 | 13,594 (89.4%) | 13,559 (89.2%) | 13,545 (89.1%) |
| Caesarean section | 3,650 | 1,698 (46.5%) | 792 (21.7%) | 792 (21.7%) |
| Birth weight | | | | |
| <1000 g | 45 | 10 (22.2%) | 9 (20.0%) | 9 (20.0%) |
| 1000–1499 g | 153 | 90 (58.8%) | 74 (48.4%) | 74 (48.4%) |
| 1500–2499 g | 2,047 | 1,517 (74.1%) | 1,352 (66.0%) | 1,351 (66.0%) |
| 2500–3999 g | 16,117 | 13,304 (82.5%) | 12,589 (78.1%) | 12,576 (78.0%) |
| ≥4000 g | 447 | 341 (76.3%) | 297 (66.4%) | 297 (66.4%) |
| Gestational age | | | | |
| <28 weeks | 39 | 13 (33.3%) | 12 (30.8%) | 12 (30.8%) |
| 28–31+6 weeks | 177 | 112 (63.3%) | 97 (54.8%) | 97 (54.8%) |

| | | | | |
|---|---|---|---|---|
| 32–36+6 weeks | 1,870 | 1,406 (75.2%) | 1,285 (68.7%) | 1,284 (68.7%) |
| ≥37 weeks | 16,753 | 13,749 (82.1%) | 12,945 (77.3%) | 12,932 (77.2%) |

**Table S4.** Characteristics of the eligible cohort (Inborn babies admitted to NICU over 12 month study period, 1 min Apgar score ≥7) with a documented DCC/ESSC status, by intervention group.

| Characteristic | Overall | Neither | DCC only | ESSC only | Both |
|---|---|---|---|---|---|
| N | 1,794 | 920 | 426 | 61 | 387 |
| Mode of delivery, n (%) | | | | | |
| Vaginal (incl. instrumental) | 1,017 (56.7%) | 483 (52.5%) | 153 (35.9%) | 54 (88.5%) | 327 (84.5%) |
| Caesarean section | 777 (43.3%) | 437 (47.5%) | 273 (64.1%) | 7 (11.5%) | 60 (15.5%) |
| Birth weight, n (%) | | | | | |
| <1000 g | 20 (1.1%) | 9 (1.0%) | 7 (1.7%) | 2 (3.3%) | 2 (0.5%) |
| 1000–1499 g | 98 (5.5%) | 53 (5.8%) | 21 (5.0%) | 6 (9.8%) | 18 (4.7%) |
| 1500–2499 g | 502 (28.0%) | 273 (29.7%) | 134 (31.6%) | 11 (18.0%) | 84 (21.7%) |
| 2500–3999 g | 998 (55.7%) | 504 (54.8%) | 215 (50.7%) | 33 (54.1%) | 246 (63.6%) |
| ≥4000 g | 174 (9.7%) | 81 (8.8%) | 47 (11.1%) | 9 (14.8%) | 37 (9.6%) |
| Gestational age, n (%) | | | | | |
| <28 weeks | 24 (1.3%) | 14 (1.5%) | 4 (0.9%) | 3 (4.9%) | 3 (0.8%) |
| 28–31+6 weeks | 88 (4.9%) | 46 (5.0%) | 20 (4.7%) | 3 (4.9%) | 19 (4.9%) |
| 32–36+6 weeks | 430 (24.0%) | 235 (25.5%) | 112 (26.4%) | 13 (21.3%) | 70 (18.1%) |
| ≥37 weeks | 1,251 (69.8%) | 625 (67.9%) | 289 (68.0%) | 42 (68.9%) | 295 (76.2%) |

*Percentages are column percentages within each group, calculated among infants with the characteristic recorded. Mode of delivery was recorded for all infants; birth weight was missing for 2 and gestational age for 1.*

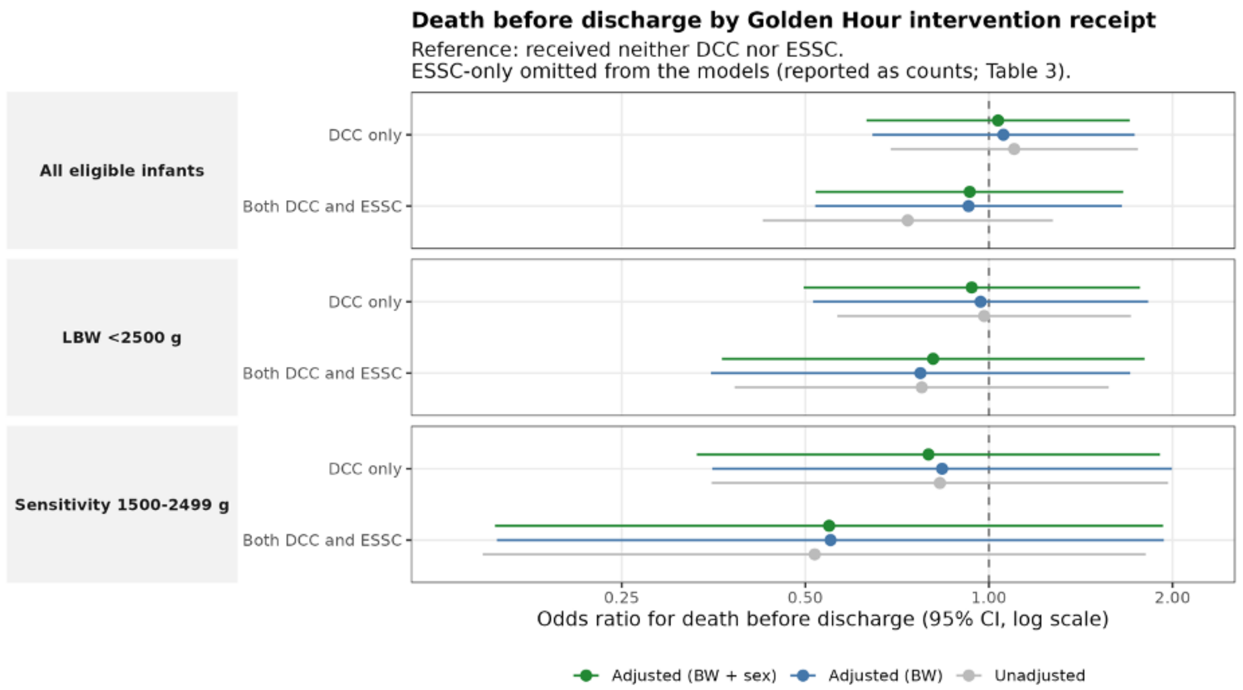


**Figure S2.** Forest plot showing odds ratios for death before discharge by Golden Hour intervention status

**Table S5.** Sensitivity analysis restricted to eligible (1min Apgar ≥7) infants weighing 1500–2499g (ESSC-only excluded)

| Intervention group | Unadjusted OR (95% CI) | Adjusted for birth weight | Adjusted for birth weight and sex |
|---|---|---|---|
| Sensitivity: 1500–2499 g (n = 456, deaths = 29) | | | |
| Neither DCC nor ESSC (reference) | Reference | Reference | Reference |
| DCC only | 0.83 (0.35–1.97) | 0.84 (0.35–2.00) | 0.80 (0.33–1.91) |
| Both DCC and ESSC | 0.52 (0.15–1.81) | 0.55 (0.16–1.94) | 0.55 (0.15–1.93) |
